# How are *APOE4*, changes in body weight, and longevity related? Insights from a causal mediation analysis

**DOI:** 10.1101/2023.12.27.23300485

**Authors:** Rachel Holmes, Hongzhe Duan, Olivia Bagley, Deqing Wu, Yury Loika, Alexander Kulminski, Anatoliy Yashin, Konstantin Arbeev, Svetlana Ukraintseva

## Abstract

The ε4 allele of the *APOE* gene (*APOE4)* is known for its negative association with human longevity, however, the mechanism is unclear. *APOE4* was also linked to changes in body weight, and the latter changes were associated with survival in some studies. Here we explore the role of aging changes in weight in the connection between *APOE4* and longevity, using a Causal Mediation Analysis (CMA) approach to uncovering mechanisms of genetic associations. Using the Health and Retirement Study (HRS) data, we tested a hypothesis, whether the association of *APOE4* with reduced survival to age 85+ is mediated by key characteristics of age-trajectories of weight, such as age at reaching peak values, and slope of the decline in weight afterwards. Mediation effects were evaluated by the Total Effect (TE), Natural Indirect Effect, and Proportion Mediated. Controlled Direct Effect and Natural Direct Effect are also reported. The CMA results suggest that *APOE4* carriers have 19%-22% (TE p=0.019-0.038) lower chances of surviving to age 85 and beyond in part because they reach peak values of weight at younger ages, and their weight declines faster afterwards, compared to non-carriers. This finding is in line with the idea that detrimental effect of *APOE4* on longevity is in part related to accelerated physical aging of ε4 carriers.

## 1 Introduction

The ε4 allele of the *APOE* gene (*APOE4)* is known for its negative association with human longevity (Gurinovich et al. 2019; Abondio et al. 2019; Sebastiani et al. 2019; Garatachea et al. 2015; Santovito, Galli, and Ruberto 2019; Kulminski et al. 2022; Arbeev et al. 2023; Kulminski et al. 2019; Yashin et al. 2018a; Yashin et al. 2018b; Zeng et al. 2016; Kulminski et al. 2014; Arbeev et al. 2012; Kulminski et al. 2011; Arbeev et al. 2011), though the mechanism is not well understood. Certain metabolic phenotypes (e.g., diabetes and serum lipids) were proposed as potential mediators, however, their mediating role in the association of *APOE4* with longevity was not confirmed (Noordam et al. 2016). *APOE4* was also linked to a lower weight and body mass index (BMI) and its changes with age (Bäckman et al. 2015; Bell et al. 2017; Blautzik et al. 2018; Ando et al. 2022; Kulminski, et al. 2019; Ukraintseva et al. 2023). E.g., we recently demonstrated using HRS and FHS data that *APOE4* carriers had lower weight than non-carriers starting around age 65, and reached maximum of the weight at younger ages compared to non-carriers, as well as declined in weight faster than non-carriers (Ukraintseva, et al. 2023).

Weight/BMI and their changes in late life were associated with survival towards older ages (Køster-Rasmussen et al. 2016; Lee et al. 2001). In a study of post-menopausal women, it was found that weight loss, especially unintentional, over a period of three to ten years, is associated with significantly lower chances of longevity (survival to ages over 90), compared to women with stable weight (Shadyab et al. 2023). Previously, we showed that entire age-trajectories of weight/BMI can significantly differ between longer- and shorter-lived individuals (Yashin et al. 2013; Yashin et al. 2016), so that the shorter-lived individuals reached maximum values of weight/BMI, and started to decline earlier in their lives, compared to the longest-livers. Trajectories of aging-changes in weight may also differ between longer- and shorter-lived strains of laboratory rodents, so that the longer-lived strains typically reach maximum values of the weight later in their lives (Turturro et al. 1999). Reaching peak values of the weight at younger ages, and faster decline in the weight afterwards, might thus be indicators of accelerated physical aging, which in turn could contribute to lower chances of extreme longevity (Yashin et al. 2010).

This and other evidence indicate a possibility that some of the *APOE4* effects on longevity may be attributed to dynamics of aging-changes in weight/BMI. Here we explore causal relationships between *APOE4* carrier status, key characteristics of mean age-trajectories of body weight (such as age at reaching maximum, and slope of decline), and survival to the oldest old age (85+), applying a Causal Mediation Analysis (CMA) (Imai, Keele, and Tingley 2010; Valeri and VanderWeele 2013) to the Health and Retirement Study (HRS) data. We propose that *APOE4* may influence age-trajectories of the weight, and the negative effect of *APOE4* on longevity could in part be because it promotes the earlier/faster decline in weight in older adults.

## 2 Materials and Methods

## 2.1 Data

For this analysis, Health and Retirement Study data was used; the HRS is sponsored by the National Institute on Aging and is conducted by the University of Michigan. Beginning in 1992, the National Institutes of Aging and the University of Michigan began collecting a variety of phenotypic information, with a focus on aging and aging related information, on participants aged 50+ in the United States. Since then, it has grown and expanded to about 20,000 participants and is now one of the largest longitudinal studies of those aged 50+ in the United States. Participants are provided with physical informed consent information prior to each interview, and are required to verbally consent. More details on the study design and sample selection can be found in Sonnega et al. (2014).

Phenotype information was derived from the RAND HRS Longitudinal File (version 2016v1) which is an easy-to-use dataset based on the HRS core data. This file was developed at RAND with funding from the National Institute on Aging and the Social Security Administration. It contains a variety of information from interviews of HRS participants, such as demographics, health, employment, and retirement.

For the present analysis, a subset of participants was selected: those who survived to at least age 80, and those with genotype information for *APOE4*. *APOE4* status was derived from imputed genotype information obtained from the database of Genotypes and Phenotypes (dbGaP) HRS data (dbGaP Study Accession: phs000428.v2.p2) which includes genotype information for 2.5 million SNPs, collected using the Omni2.5 BeadChip. Further details regarding characteristics of participants in the entire HRS sample and in the analytic sample can be found in Tables 1a and 1b.

**Table 1a:** Characteristics of entire HRS sample.

| HRS | Males |  | Females |  | Total |  |
| --- | --- | --- | --- | --- | --- | --- |
| *APOE4* Carrier | No | Yes | No | Yes | No | Yes |
| N | 4674 | 1678 | 6545 | 2430 | 11219 | 4108 |
| **Continuous Variables: Mean (Standard Deviation)** |  |  |  |  |  |  |
| *Entry Age* | 57.86(7.82) | 57.64(7.20) | 56.68(9.21) | 56.17(8.65) | 57.17(8.68) | 56.77(8.39) |
| *End of Follow-Up Age* | 75.33(10.44) | 75.01(10.13) | 75.14(11.44) | 74.52(10.96) | 75.22(11.04) | 74.72(10.63) |
| *Birth Cohort* | 1940.68(11.38) | 1940.82(11.08) | 1941.26(12.20) | 1941.89(11.72) | 1941.02(11.87) | 1941.45(11.47) |
| **Dichotomous Variables: N(percentage): number and percentage of individuals in group '1'.** |  |  |  |  |  |  |
| *Education, Highschool+* | 4166(89.38) | 1448(89.00) | 5902(90.41) | 2212(91.33) | 10068(89.98) | 3700(90.38) |
| *Ever Smoked* | 3132(67.41) | 1135(68.25) | 3139(48.24) | 1225(50.72) | 6271(56.23) | 2360(57.87) |
| *AgeMaxBMI* | 696(46.12) | 221(41.54) | 1089(51.91) | 316(42.70) | 1785(49.49) | 537(42.22) |
| *SlopeBMI* | 711(51.75) | 216(44.35) | 983(52.40) | 296(43.72) | 1694(52.12) | 512(43.99) |
| *AgeMaxW* | 562(37.24) | 179(33.65) | 848(40.57) | 239(32.34) | 1410(39.18) | 418(32.89) |
| *SlopeW* | 705(51.31) | 225(46.20) | 990(52.83) | 292(43.00) | 1695(52.19) | 517(44.34) |
| *Survival85* | 929(53.27) | 315(47.73) | 1411(64.84) | 444(57.66) | 2340(59.69) | 759(53.08) |
| *Race: Black* | 545(11.67) | 319(19.02) | 945(14.45) | 538(22.15) | 1490(13.29) | 857(20.87) |
| *Race: White* | 3442(73.69) | 1182(70.48) | 4599(70.30) | 1619(66.65) | 8041(71.71) | 2801(68.22) |

### 2.2 Statistical Analysis

In general, mediation analysis concerns two main pathways: direct and indirect. Illustrated by Figure 1, the direct pathway is seen by the effect of the “treatment” (*APOE4* carrier status in our case) on the outcome. The indirect path is illustrated by the indirect effect of the “treatment” on the outcome, as mediated through the mediator.

**Figure 1.**
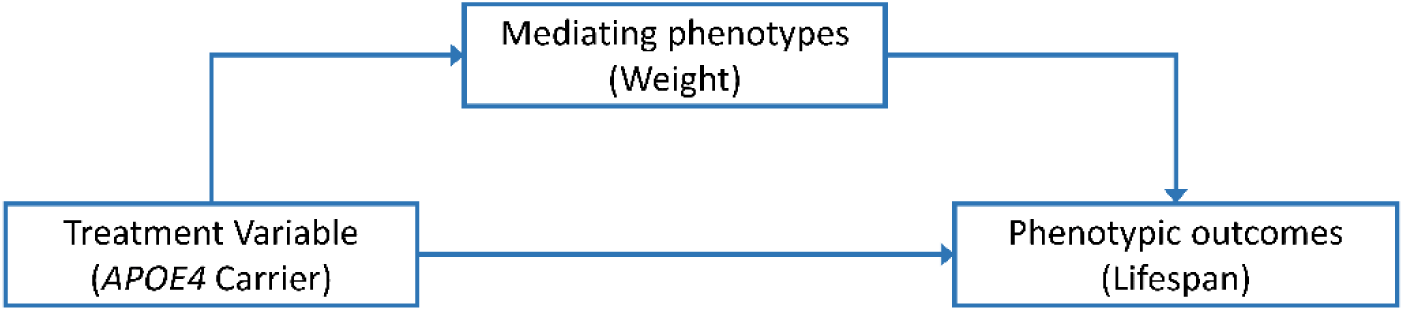
A simplified description of the idea behind a Causal Mediation Analysis (CMA)

Mediation analysis measures the direct and indirect effects illustrated in Figure 1, based on an outcome model and a mediator model. The general form of these models, respectively, is:

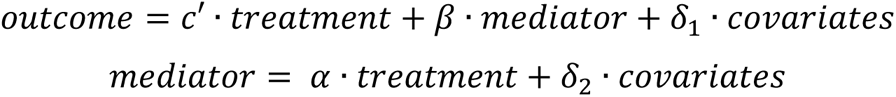

Specifically, direct effect is measured by the effect of treatment on outcome, 𝑐′, and indirect effect is measured by 𝛼𝛽. That is, the total effect, commonly denoted as 𝑐, can be expressed by

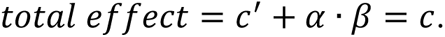

Additionally, there exists a measure to determine amount of mediation, called the Percentage Mediated (PM), which is calculated by dividing the indirect effect by the total effect:

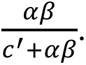

This measures the percentage of the effect of the treatment on the outcome which can be attributed to the mediator. In order to claim “complete mediation”, the PM should be at least 80% (Kenny 2021).

We performed Causal Mediation Analysis, an extension of Traditional Mediation Analysis (TMA) (Baron and Kenny 1986), in the HRS data analytic sample (see Table 1b), using the SAS software Version 9.4. CMA utilizes a counterfactual framework to describe direct, indirect, and total effect estimates using natural and controlled effects. In this analysis, for example, a counterfactual approach consists of evaluating the chances of survival for each individual, at both “treatment” levels, even though each individual can only have one “treatment” level (i.e., and individual cannot be a carrier and a non-carrier of *APOE4*). The potential outcome for all individuals is then averaged to calculate the overall potential outcome at each “treatment” level. A variety of potential outcomes at differing mediator and “treatment” levels is used to calculate specific CMA estimates. Additionally, CMA effects are not bound by parametric assumptions, unlike TMA effects (Rijnhart et al. 2021).

**Table 1b:** Characteristics of HRS Participants in *APOE4* subsample who lived to age ≥80.

| HRS | Males |  | Females |  | Total |  |
| --- | --- | --- | --- | --- | --- | --- |
| *APOE4* Carrier | No | Yes | No | Yes | No | Yes |
| N | 1655 | 576 | 2331 | 805 | 3986 | 1381 |
| **Continuous Variables: Mean (Standard Deviation)** |  |  |  |  |  |  |
| *Entry Age* | 65.41(7.38) | 65.22(7.32) | 65.89(7.47) | 65.13(7.28) | 65.69(7.44) | 65.17(7.30) |
| *End of Follow-Up Age* | 86.60(4.72) | 86.12(4.44) | 87.48(5.34) | 86.76(5.06) | 87.11(5.11) | 86.49(4.82) |
| *Birth Cohort* | 1928.69(5.97) | 1929.08(5.63) | 1928.32(6.38) | 1928.95(6.17) | 1928.48(6.21) | 1929.00(5.95) |
| **Dichotomous Variables: N(percentage): number and percentage of individuals in group '1'.** |  |  |  |  |  |  |
| *Education, Highschool+* | 1446(87.37) | 488(84.87) | 2064(88.55) | 722(89.69) | 3510(88.06) | 1210(87.68) |
| *Ever Smoked* | 1121(68.52) | 401(70.47) | 1001(43.17) | 354(44.19) | 2122(53.65) | 755(55.11) |
| *AgeMaxBMI* | 696(46.12) | 221(41.54) | 1089(51.91) | 316(42.70) | 1785(49.49) | 537(42.22) |
| *SlopeBMI* | 711(51.75) | 216(44.35) | 983(52.40) | 296(43.72) | 1694(52.12) | 512(43.99) |
| *AgeMaxW* | 562(37.24) | 179(33.65) | 848(40.57) | 239(32.34) | 1410(39.18) | 418(32.89) |
| *SlopeW* | 705(51.31) | 225(46.20) | 990(52.83) | 292(43.00) | 1695(52.19) | 517(44.34) |
| *Survival85* | 929(79.40) | 315(76.64) | 1411(86.67) | 444(81.02) | 2340(83.63) | 759(79.14) |
| *Race: Black* | 143(8.64) | 65(11.28) | 239(10.25) | 125(15.53) | 382(9.58) | 190(13.76) |
| *Race: White* | 1374(83.02) | 466(80.90) | 1868(80.14) | 624(77.52) | 3242(81.33) | 1090(78.93) |

*APOE4* status was used as a binary “treatment” variable in the CMA: (1) *APOE4* carriers (“treatment” group) vs. (0) *APOE4* non-carriers (control group). The longevity-related outcome, *Survival85*, was a binary variable defined as: (1) survival to age 85 years and above, vs. (0) died before age 85. Two mediator variables characterizing weight were considered. The first variable, *AgeMaxW*, was defined as (1) age at maximum weight ≥75 years vs. (0) age at maximum weight <75 years, and age at last follow up or death >80 years. The second variable, *SlopeW* was defined as (1) relative weight change above median (tend to have more stable weight during ages 65-80) vs. (0) relative weight change below median (tend to be losing weight during ages 65-80). The *SlopeW* was calculated as: (mean value at ages 75-79 - mean value at ages 65-74)/(mean value at ages 65-74), and dichotomized among individuals with age at last follow up or death > 80 years.

The SAS procedure PROC CAUSALMED (SAS Institute Inc. 2021) with the SAS software Version 9.4 was used for the entire analysis. As the outcome and mediators are binary variables, logistic regression models were used for the CMA. The trust-region optimization technique (‘TRUREG’ option) was used to obtain maximum likelihood estimates. The codes used to generate the reported output are available upon request from the corresponding author.

For our CMA approach, we focused primarily on three causal effect estimates: Total Effect (TE), Natural Indirect Effect (NIE), and Proportion Mediated (PM). In general, TE, a combination of both the direct and indirect effects, provides an estimate of “treatment” effect on the outcome at the various mediator levels. Here, this estimate compares the chances of survival for *APOE4* carriers vs. non-carriers, while taking into account the effect of the mediator. NIE provides further details about the effects of “treatment” through the mediator, by holding the treatment constant and considering the effect of the mediator only. In this analysis, this estimate will compare chances of survival for allele carriers at each of the two mediator levels. PM gives the percentage of the total effect that is mediated by the mediator. Other reported estimates include Natural Direct Effect (NDE) and Controlled Direct Effect (CDE). While both of these estimates are useful, they were not the focus of this study as they hold the effect of the mediator constant, i.e., these estimates compare the chances of survival for the two “treatment” groups (carriers vs. non-carriers), but without considering the effect of the mediator (SAS Institute Inc. 2021).

The CMA approach has four assumptions (VanderWeele 2016): control must be made for (Assumption 1) exposure-outcome confounding, (Assumption 2) mediator-outcome confounding, and (Assumption 3) exposure-mediator confounding, as well as (Assumption 4) no mediator-outcome confounders are affected by the treatment. The following covariates were included in both the mediator and outcome model in order to control for confounding: smoking status (coded 0 – never smoked, 1 – ever smoked), education (coded 0 – less than high school, 1 – high school only, 2 – higher education), sex (coded 1 – male, 2 – female), race (coded 1 – White, 2 – Black, 3 – Other). Furthermore, we followed A Guideline for Reporting Mediation Analyses (AGReMA) long-form checklist (Lee et al. 2021) (see Table S1). See (VanderWeele 2015) for additional details and methods.

In addition to reporting results for the total analytic sample, we provided stratified results by sex and race. Instead of creating sex and race subsamples, the ‘Evaluate’ statement was used within PROC CAUSALMED. This allows users to specify covariate levels (such as sex and race) and evaluate respective conditional causal effects. The ‘Evaluate’ statement uses the entire analytic sample and estimates from the associated model to compute the mean of the specified covariate level(s). SAS documentation on PROC CAUSALMED provides further details (SAS Institute Inc. 2021). Additionally, we only used observations for which all variables involved in either model are non-missing (i.e., only those observations with information for “treatment,” outcome, mediator, and all covariates were used).

Furthermore, using the HRS data, we compared mean trajectories of the weight for *APOE4* carriers and non-carriers, as well as survival trajectories by different weight slopes and ages at reaching maximum values of weight (Figure 2). These variables are proxies to respective variables defined for BMI because height does not generally demonstrate substantial changes in individuals at ages over 80. Further, a Cox Proportional Hazards p-value is provided on each plot as a measure of significance of survival models.

**Figure 2.**
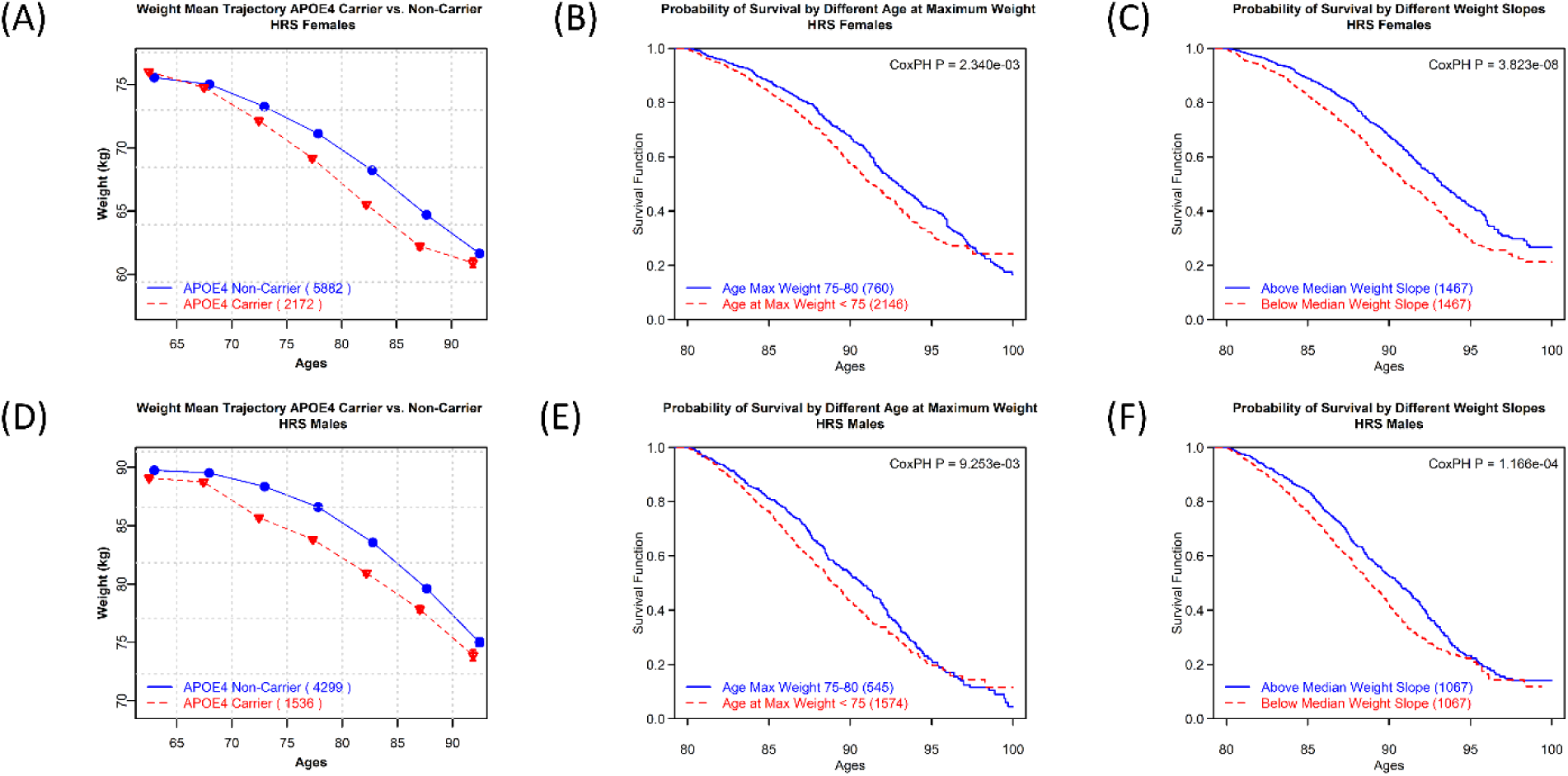
**HRS Females: A.** Mean trajectories of body weight in *APOE4* carriers vs. non-carriers. **B**. Survival trajectories by different age at reaching maximum values of the weight. **C**. Survival trajectories by different weight slopes. **HRS Males: D.** Mean trajectories of body weight in *APOE4* carriers vs. non-carriers. **E**. Survival trajectories by different age at reaching maximum values of the weight. **F**. Survival trajectories by different weight slopes. Cox Proportional Hazards p-value (CoxPH P) is provided as a measure of statistical significance of differences between survival curves in respective groups.

### 2.3 Sensitivity Analysis

Since the Ɛ2 allele of the *APOE* gene is known for its beneficial effects on longevity (Wolters et al. 2019), we also conducted a sensitivity analysis with this allele removed. That is, we compared those with Ɛ3/Ɛ3 genotype (non-carriers) against those with Ɛ3/Ɛ4 or Ɛ4/Ɛ4 genotypes (carriers).

## 3 Results

Mean trajectories of the weight for *APOE4* carriers and non-carriers, as well as survival trajectories by different weight slopes and different age at reaching maximum values of the weight are shown in Figure 2. These variables are proxies for similarly defined BMI variables, as height does not show substantial changes for those age 80+. *APOE4* carriers decline faster in weight with age, compared to non-carriers (see Figure 2.A, 2.D). Survival was better for individuals with less steep weight slope, and older age at reaching maximum values of the weight (after age 75 vs less than 75 years) (see Figures 2.B, 2.C, 2.E, 2.F).

***AgeMaxW*** (below 75 years) was a significant mediator of the effects of the *APOE* alleles on survival to ages 85+ in the total sample, and in samples stratified by sex and race. This was supported by the significant p-values for TE and NIE (see Table 2). TE estimates suggest that ε4 carriers are 19%-22% (p=0.019-0.038) more likely to die before age 85, compared to non-carriers. If we hold the “treatment” constant and consider only the mediator effect, then *APOE4* carriers who have a younger *AgeMaxW* are about 2.5% (p=0.027-0.029) more likely to die before age 85 than the carriers who have an older *AgeMaxW*. The percentage of the total effect mediated by the *AgeMaxW* was around 14% for the overall sample and all strata, with p=0.074-0.075.

**Table 2.** Results of CMA for AgeMaxW and SlopeW, evaluated at different strata, for survival 85+. (TE [LC, UC], pval): (Total Effect estimate [Lower 95% Confidence interval, Upper 95% Confidence interval], p-value of Total Effect estimate), similarly for NIE (Natural Indirect Effect), PM (Proportion Mediated), CDE (Controlled Direct Effect), and NDE (Natural Direct Effect). Effect estimates are presented on the Relative Risk scale. **\*=s**ignificant p-value (<0.05)

| Mediator | Strata | (TE [LC,UC], pval) | (NIE [LC,UC], pval) | (PM [LC,UC], pval) | (CDE [LC,UC], pval) | (NDE [LC,UC], pval) |
| --- | --- | --- | --- | --- | --- | --- |
| AgeMaxW | Total | (1.215 [1.036, 1.394], 0.019)* | (1.025 [1.003, 1.048], 0.029)* | (14.033 [-1.365, 29.430], 0.074) | (1.473 [1.062, 1.884], 0.024)* | (1.185 [1.009, 1.361], 0.039)* |
| AgeMaxW | Male | (1.206 [1.029, 1.384], 0.023)* | (1.025 [1.003, 1.047], 0.028)* | (14.189 [-1.351, 29.730], 0.074) | (1.473 [1.062, 1.884], 0.024)* | (1.177 [1.003, 1.351], 0.046)* |
| AgeMaxW | Female | (1.211 [1.033, 1.390], 0.020)* | (1.025 [1.003, 1.048], 0.028)* | (14.104 [-1.357, 29.565], 0.074) | (1.473 [1.062, 1.884], 0.024)* | (1.182 [1.006, 1.357], 0.042)* |
| AgeMaxW | White | (1.211 [1.033, 1.390], 0.020)* | (1.025 [1.003,1.048], 0.028)* | (14.104 [-1.357, 29.565], 0.074) | (1.473 [1.062, 1.884], 0.024)* | (1.182 [1.006, 1.357], 0.042)* |
| AgeMaxW | Black | (1.186 [1.010, 1.362], 0.038)* | (1.024 [1.003, 1.044], 0.027)* | (14.404 [-1.448, 30.356], 0.075) | (1.473 [1.062, 1.884], 0.024)* | (1.156 [0.989, 1.330], 0.067) |
| SlopeW | Total | (1.224 [1.036, 1.413], 0.020)* | (1.036 [1.007, 1.065], 0.015)* | (18.889 [-0.469, 38.248], 0.056) | (1.258 [0.936, 1.580], 0.117) | (1.182 [0.998, 1.366], 0.053) |
| SlopeW | Male | (1.223 [1.035, 1.411], 0.020)* | (1.036 [1.007, 1.064], 0.015)* | (18.914 [-0.388, 38.216], 0.055) | (1.258 [0.936, 1.580], 0.117) | (1.181 [0.997, 1.364], 0.054) |
| SlopeW | Female | (1.222 [1.034, 1.411], 0.021)* | (1.036 [1.007, 1.064], 0.015)* | (18.925 [-0.344, 38.194], 0.054) | (1.258 [0.936, 1.580], 0.117) | (1.180 [0.997, 1.364], 0.054) |
| SlopeW | White | (1.222 [1.034, 1.411], 0.021)* | (1.036 [1.007, 1.064], 0.015)* | (18.925 [-0.344, 38.194], 0.054) | (1.258 [0.936, 1.580], 0.117) | (1.180 [0.997, 1.364], 0.054) |
| SlopeW | Black | (1.223 [1.034, 1.411], 0.021)* | (1.036 [1.007, 1.064], 0.015)* | (18.921 [-0.357, 38.199], 0.054) | (1.258 [0.936, 1.580], 0.117) | (1.181 [0.997, 1.364], 0.054) |

***SlopeW*** was also a significant mediator of the *APOE4* effects on survival 85+ in the total sample, with about a 4%-higher (NIE: p-value=0.015) (see Table 2) chance of death before age 85 when holding the “treatment” constant and considering the mediator’s effect only. TE estimates suggested chances of dying before age 85 were about 22% higher for *APOE4* carriers vs. non-carriers (p-value = 0.020-0.021). A very similar effect was seen at all strata levels, with similar p-values. The percentage of the total effect mediated by *SlopeW*, around 19%, was nearly significant (p=0.054-0.056).

In our sensitivity analysis, which has the same framework as initial analysis, but with 𝜀2 carriers removed from *APOE4* “treatment”, results of both *AgeMaxW* and *SlopeW* are comparable to initial results and are available in Supplementary Table S2. It is worth noting that these results are “improved” slightly, in terms of estimated size and significance for TE and NIE, and similar in size and effect for other estimates.

## 4 Discussion

Findings of this CMA suggest that dynamics of aging-changes in body weight mediates the effect of *APOE4* on longevity. Specifically, *APOE4* carriers have lower chances of survival to age 85+, and estimates suggest that 14-22% of this association is related to reaching maximum of the weight at earlier ages. The *APOE4* carriers were about 19-22% more likely to die before age 85, compared to non-carriers, when considering the mediating effects of *SlopeW* and *AgeMaxW*. To our knowledge, no studies have yet considered patterns of aging changes in body weight over the life course as mediators of the negative association between *APOE4* and longevity.

One may argue that the well-known beneficial effects of the Ɛ2 allele of the *APOE* gene (Wolters, et al. 2019) may contribute to the difference in survival chances seen in this CMA. To account for this, and to ensure that *APOE4’*s effect on longevity is indeed mediated by aging-changes in body weight, we conducted a sensitivity analysis with Ɛ2 allele carriers removed from the “treatment” variable. This increased the effect size and significance of most TE and NIE estimates (Supplementary Table S2). Other estimates are relatively similar in effect size and significance, though still vary from the original CMA results. This further suggests that the effect of *APOE4* on longevity is partially mediated by aging-changes in body weight, and the Ɛ2 allele is not the source of the difference in survival chances seen in this CMA.

One should emphasize that the CMA that was implemented in this study has advantages over TMA. Since TMA effects are based on coefficients of regression models, it is bound by parametric assumptions, whereas natural effects from CMA are not. Additionally, TMA does not usually include an interaction between treatment and mediator (Rijnhart, et al. 2021). Another advantage is the use of a causal inference approach which utilizes a counterfactual framework. While TMA is a useful tool for detecting mediating effects, and describing total, direct, and indirect effects, CMA breaks these effects down even more, helping to better understand the underlying mechanism of the negative association between *APOE4* and longevity.

Our results are in line with earlier studies that demonstrated a detrimental effect of *APOE4* on longevity (Gurinovich, et al. 2019; Abondio, et al. 2019; Sebastiani, et al. 2019; Garatachea, et al. 2015; Santovito, Galli, and Ruberto 2019; Arbeev, et al. 2011; Kulminski, et al. 2022; Kulminski, et al. 2019; Yashin, et al. 2018a; Yashin, et al. 2018b; Zeng, et al. 2016; Kulminski, et al. 2014; Arbeev, et al. 2012; Kulminski, et al. 2011). They also suggest that *APOE4* facilitates physical aging (manifested in earlier and faster decline in the weight) (Yashin, et al. 2010), which in turn can causally contribute to reduced longevity. Previously, we showed that longest-lived individuals reach peak values of weight/BMI and start to decline later in their lives, as compared to people with conventional lifespan (Yashin, et al. 2013; Yashin, et al. 2016). The ability to grow in size and postpone weight loss till older ages may also correlate with better physical resilience to life stressors, which is essential for survival at the oldest old ages (Ukraintseva et al. 2021).

A potential limitation of this study is that, while highly relevant covariates were included in both models (smoking status, education level, sex, and race), there still may exist unmeasured confounders, creating the potential for biased effect estimates. This CMA also revealed that *AgeMaxW* and *SlopeW* are factors that may explain only a part (albeit substantial) of the negative association between *APOE4* and longevity, because they did not meet the criteria to satisfy ‘complete mediation’ (𝑃𝑀 ≥ 80%) (Kenny 2021). This suggests that there are other causal factors to be discovered, which warrants further investigation of the mechanisms involved in the association between *APOE4* and longevity, using other mediators, such as those involved in dysregulations of cholesterol transport, impaired myelination, hypometabolism, and other traits associated with *APOE4* that may also affect longevity (Zhang et al. 2023; Blanchard et al. 2022).

In conclusion, this Causal Mediation Analysis found that *APOE4* carriers have lower chances of surviving to age 85 and beyond in part because they reach peak values of body weight at younger ages, and decline faster afterwards, compared to non-carriers. Postponing physical aging in *APOE4* carriers could be a promising target of pro-longevity interventions.

## 5 Ethics Statement

This study involves secondary analyses of existing human data and was approved by the Duke University Health System IRB.

## 7 Author Contributions

SU conceived the study, developed research hypothesis, provided interpretation of results, and wrote the manuscript. RH designed the study, conducted statistical analyses, and wrote the manuscript. HD, KA, and AY supervised statistical analyses and data preparation, and contributed to writing of the manuscript and discussion of its content. AK, DW, and YL contributed to discussion of the manuscript content. OB contributed to data preparation. YL assisted in genetic data imputation. All authors read and approved the final manuscript.

## 9 Acknowledgments

The Health and Retirement Study (HRS) is sponsored by the National Institute on Aging (grant number U01AG009740) and conducted by the University of Michigan. The HRS genetic data was sponsored by the National Institute on Aging (grants U01AG009740, RC2AG036495, and RC4AG039029) and collected by the University of Michigan. Access to the HRS genetic data is provided by the database of Genotypes and Phenotypes, dbGaP (Study Accession: phs000428.v2.p2).

## Data Availability Statement

The datasets presented in this article are not readily available because of dbGaP and the University of Michigan Data Use Certification Agreement restrictions. Requests to access the datasets should be directed to the RAND HRS data provider.

