## Supplemental Tables for "How are *APOE4*, changes in body weight, and longevity related? Insights from a causal mediation analysis"

Supplementary Material

### 1 Supplementary Tables

**Table S1.**  A Guideline for Reporting Mediation Analyses (AGReMA) long-form checklist (Lee, et al. 2021).

| **Section and Topic** | **Item No.** | **Item description** | **Location** |
| --- | --- | --- | --- |
| **Title and Abstract** |  |  |  |
| Title | 1 | Identify that the study uses mediation analyses | p.1 |
| Abstract | 2 | Provide a structured summary of the objectives, methods, results, and conclusions specific to mediation analyses | p.1 |
| **Introduction** |  |  |  |
| Background and Rationale | 3 | Describe the study background and theoretical rationale for investigating the mechanisms of interest | 1 |
|  | 4 | Include supporting evidence or theoretical rationale for why the intervention or exposure might have a causal relationship with the proposed mediators | 1 |
|  | 5 | Include supporting evidence or theoretical rationale for why the mediators might have a causal relationship with the outcomes | 1 |
| Objectives | 6 | State the objectives of the study specific to the mechanisms of interest  The objectives should specify whether the study aims to test or estimate the mechanistic effects | 1 |
| **Methods** |  |  |  |
| Study Registration | 7 | If applicable, provide references to any protocols or study registrations specific to mediation analyses and highlight any deviations from the planned protocol | N/A |
| Study Design and Source of Data | 8 | Specify the design of the original study that was used in mediation analyses and where the details can be accessed, supported by a reference | 2.1 |
|  | 9 | If applicable, describe study design features that are relevant to mediation analyses | N/A |
| Participants | 10 | Describe the target population, eligibility criteria specific to mediation analyses, study locations, and study dates (start of participant enrollment and end of follow-up) | 2.1 |
| Sample Size | 11 | State whether a sample size calculation was conducted for mediation analyses | 2.2 |
|  | 12 | If so, explain how it was calculated | 2.2 |
| Effects of Interest | 13 | Specify the effects of interest | 2.2 |
| Assumed Causal Model | 14 | Include a graphic representation of the assumed causal model including the exposure, mediator, outcome, and possible confounders | Figure 1 |
| Causal Assumptions | 15 | Specify assumptions about the causal mode | 2.2 |
| Measurement | 16 | Clearly describe the interventions or exposures, mediators, outcomes, confounders, and moderators that were used in the analyses | 2.2 |
|  | 17 | Specify how and when they were measured, the measurement properties, and whether blinded assessment was used | 2.2 |
| Measurement Levels | 18 | If relevant, describe the levels at which the exposure, mediator, and outcome were measured | 2.2 |
| Statistical Methods | 19 | Describe the statistical methods used to estimate the causal relationships of interest | 2.2 |
|  | 20 | This description should specify analytic strategies used to reduce confounding, model building procedures, justification for the inclusion or exclusion of possible interaction terms, modeling assumptions, and methods used to handle missing data | 2.2 |
|  | 21 | Provide a reference to the statistical software and package used | 2.2 |
| Sensitivity Analyses | 22 | Describe any sensitivity analyses that were used to explore causal or statistical assumptions and the influence of missing data | 2.3 |
| Ethical Approval | 23 | Name the institutional research board or ethics committee that approved the study | 5 |
|  | 24 | Provide a description of participant informed consent or ethics committee waiver of informed consent N/A | 2.1 |
| **Results** |  |  |  |
| Participants | 25 | Describe baseline characteristics of participants included in mediation analyses | Table 1 |
|  | 26 | Report the total sample size and number of participants lost during follow-up or with missing data | **Table 2** |
| Outcomes and Estimates | 27 | Report point estimates and uncertainty estimates for the exposure-mediator and mediator-outcome relationships | **Table 2** |
|  | 28 | If inference concerning the causal relationship of interest is considered feasible given the causal assumptions, report the point estimate and uncertainty estimate | NA |
| Sensitivity Parameters | 29 | Report the results from any sensitivity analyses used to assess robustness of the causal or statistical assumptions and the influence of missing data | Table S2 |
| **Discussion** |  |  |  |
| Limitations | 30 | Discuss the limitations of the study including potential sources of bias | 4 |
| Interpretation | 31 | Interpret the estimated effects considering the study’s magnitude and uncertainty, plausibility of the causal assumptions, limitations, generalizability of the findings, and results from relevant studies | 4 |
| Implication | 32 | Discuss the implications of the overall results for clinical practice, policy, and science | 4 |
| **Other Information** |  |  |  |
| Funding and Role of Sponsor | 33 | List all sources of funding or sponsorship for mediation analyses and the role of the funders/sponsors in the conduct of the study, writing of the manuscript, and decision to submit the manuscript for publication | 8, Acknowledgments |
| Conflicts of Interest and Financial Disclosure | 34 | State any conflicts of interest and financial disclosures for all authors | 6 |
| Data and Code | 35 | Authors are encouraged to provide a statement for sharing data and code for mediation analyses | 2.2, Data Availability Statement |

**Table S2.** Results of sensitivity analysis of CMA for AgeMaxW and SlopeW, evaluated at different strata, for survival 85+, where “treatment” (*APOE4* Carrier vs. Non-Carrier) excludes the beneficial ε2 allele. (TE [LC, UC], pval): (Total Effect estimate [Lower 95% Confidence interval, Upper 95% Confidence interval], p-value of Total Effect estimate), similarly for NIE (Natural Indirect Effect), PM (Proportion Mediated), CDE (Controlled Direct Effect), and NDE (Natural Diirect Effect). Effect estimates are presented on the Relative Risk scale. ***=s**ignificant p-value (<0.05)

| **Mediator** | **Strata** | **(TE [LC,UC], pval)** | **(NIE [LC,UC], pval)** | **(PM [LC,UC], pval)** | **(CDE [LC,UC], pval)** | **(NDE [LC,UC], pval)** | **NobsUsed** |
| --- | --- | --- | --- | --- | --- | --- | --- |
| AgeMaxW | Total | (1.238 [1.042, 1.435], 0.017)* | (1.030 [1.004, 1.056], 0.024)* | (15.208 [-0.912, 31.329], 0.065) | (1.397 [0.975, 1.819], 0.065) | (1.202 [1.010, 1.395], 0.039)* | 2758 |
| AgeMaxW | Male | (1.233 [1.037, 1.428], 0.020)* | (1.030 [1.004, 1.055], 0.023)* | (15.193 [-0.815, 31.202], 0.063) | (1.397 [0.975, 1.819], 0.065) | (1.197 [1.007, 1.388], 0.042)* | 2758 |
| AgeMaxW | Female | (1.235 [1.041, 1.434], 0.018)* | (1.030 [1.004, 1.056], 0.024)* | (15.208 [-0.890, 31.307], 0.064) | (1.397 [0.975, 1.819], 0.065) | (1.201 [1.009, 1.393], 0.040)* | 2758 |
| AgeMaxW | White | (1.237 [1.041, 1.434], 0.018)* | (1.030 [1.004, 1.056], 0.024)* | (15.208 [-0.890, 31.307], 0.064) | (1.397 [0.975, 1.819], 0.065) | (1.201 [1.009, 1.393], 0.040)* | 2758 |
| AgeMaxW | Black | (1.218 [1.024, 1.411], 0.028)* | (1.028 [1.004, 1.051], 0.023)* | (14.958 [-0.677, 30.592], 0.061) | (1.397 [0.975, 1.819], 0.065) | (1.185 [0.998, 1.372], 0.053) | 2758 |
| SlopeW | Total | (1.247 [1.039, 1.455], 0.020)* | (1.042 [1.009,1.074], 0.012)* | (20.296 [0.222, 40.370], 0.048)* | (1.187 [0.856, 1.518], 0.269) | (1.197 [0.995, 1.399], 0.056) | 2521 |
| SlopeW | Male | (1.247 [1.039, 1.455], 0.020)* | (1.042 [1.009,1.074], 0.012)* | (20.310 [0.192, 40.429], 0.048)* | (1.187 [0.856, 1.518], 0.269) | (1.197 [0.995, 1.399], 0.057) | 2521 |
| SlopeW | Female | (1.247 [1.039, 1.455], 0.020)* | (1.042 [1.009,1.074], 0.012)* | (20.267 [0.281, 40.253], 0.047)* | (1.187 [0.856, 1.518], 0.269) | (1.197 [0.995, 1.399], 0.056) | 2521 |
| SlopeW | White | (1.247 [1.039, 1.455], 0.020)* | (1.042 [1.009,1.074], 0.012)* | (20.267 [0.281, 40.253], 0.047)* | (1.187 [0.856, 1.518], 0.269) | (1.197 [0.995, 1.399], 0.056) | 2521 |
| SlopeW | Black | (1.247 [1.039, 1.455], 0.020)* | (1.042 [1.009,1.074], 0.012)* | (20.282 [0.230, 40.334], 0.047)* | (1.187 [0.856, 1.518], 0.269) | (1.197 [0.995, 1.399], 0.056) | 2521 |
